# Surgically Implanted Electrodes Enable Real-Time Finger and Grasp Pattern Recognition for Prosthetic Hands

**DOI:** 10.1101/2020.10.28.20217273

**Authors:** A. K. Vaskov, P. P. Vu, N. North, A. J. Davis, T. A. Kung, D. H. Gates, P. S. Cederna, C. A. Chestek

**Affiliations:** Robotics Institute, University of Michigan, Ann Arbor, MI 48109, USA; Department of Biomedical Engineering, University of Michigan, Ann Arbor, MI 48109, USA; Department of Mechanical Engineering, University of Michigan, Ann Arbor, MI 48109, USA; University of Michigan Hospital Orthotics and Prosthetics Center, Ann Arbor, MI 48104, USA; Department of Physical Medicine and Rehabilitation, University of Michigan, Ann Arbor, MI 48109, USA; Section of Plastic Surgery, University of Michigan, Ann Arbor, MI 48109, USA; School of Kinesiology, University of Michigan, Ann Arbor, MI 48109, USA; Department of Electrical Engineering and Computer Science, University of Michigan, Ann Arbor, MI 48109, USA; Neuroscience Graduate Program, University of Michigan, Ann Arbor, MI 48109, USA

**Author notes:** corresponding author. (C.A.C).

## Abstract

Currently available prosthetic hands are capable of actuating anywhere from five to 30 degrees of freedom (DOF). However, grasp control of these devices remains unintuitive and cumbersome. To address this issue, we propose directly extracting finger commands from the neuromuscular system via electrodes implanted in residual innervated muscles and regenerative peripheral nerve interfaces (RPNIs). Two persons with transradial amputations had RPNIs created by suturing autologous free muscle grafts to their transected median, ulnar, and dorsal radial sensory nerves. Bipolar electrodes were surgically implanted into their ulnar and median RPNIs and into their residual innervated muscles. The implanted electrodes recorded local electromyography (EMG) with Signal-to-Noise Ratios ranging from 23 to 350 measured across various movements. In a series of single-day experiments, participants used a high speed pattern recognition system to control a virtual prosthetic hand in real-time. Both participants were able to transition between 10 pseudo-randomly cued individual finger and wrist postures in the virtual environment with an average online accuracy of 86.5% and latency of 255 ms. When the set was reduced to five grasp postures, average metrics improved to 97.9% online accuracy and 135 ms latency. Virtual task performance remained stable across untrained static arm positions while supporting the weight of the prosthesis. Participants also used the high speed classifier to switch between robotic prosthetic grips and complete a functional performance assessment. These results demonstrate that pattern recognition systems can use the high-quality EMG afforded by intramuscular electrodes and RPNIs to provide users with fast and accurate grasp control.

**Summary:** Surgically implanted electrodes recorded finger-specific electromyography enabling reliable finger and grasp control of an upper limb prosthesis.

## Introduction

Hands are incredibly important to people because they provide the opportunity to handle tools, operate machines, and are essential components of social interaction and communication. The loss of an upper extremity can severely impact a person’s ability to interact with the world around them. Unsurprisingly, some of the earliest documented examples of upper extremity prostheses produced in the sixteenth century mimicked the human hand. However, these devices were heavy, lacked active actuation, and were not practical for use in daily life (*1*). In the twentieth century, the development of the split hook body powered prosthesis provided a more functional solution that addressed many of these concerns (*2*). Body-powered hook prostheses are cable-driven devices that are controlled by exaggerated movements of residual joints. Unfortunately, they have very limited range of motion limiting their capacity to provide functional restoration (*3*–*5*). Myoelectric prosthetic hooks can mitigate some of these issues but still lack dexterity and fine motor control. Advanced robotic prosthetic devices have been developed which can provide 5 to 30 degrees of freedom (DOF) of the human hand and provide adequate gripping force for functional tasks (*6*–*8*). Control of these devices is achieved by recording surface electromyography (EMG) signals from innervated muscles in the residual limb, allowing users to leverage more natural motor pathways to control the device. However, current control schemes are either cumbersome, unintuitive, or unreliable, leading many users to abandon their devices (*9*). In spite of these issues, myoelectric hands are still an attractive approach for prosthetic rehabilitation following limb loss due to their high grip strength and potential for intuitive control (*10, 11*).

Commercial EMG control systems typically rely on a mode selection scheme, where the user first selects between different movements and then secondarily activates them. In systems with an active wrist, users typically toggle between wrist and hand control with a pre-defined muscle contraction. Grasp selection is often done sequentially with muscle co-contraction or gesture control (*12*). A more fluid control strategy is pattern recognition in which the intended movement is classified from the users’ recorded EMG signals and can be proportionally activated based on signal amplitude (*13*). Most pattern recognition systems can independently activate wrist rotation, wrist flexion, or a general hand open-close signal. Grasp distinction is not common, even amongst persons with transradial amputations where extrinsic flexors and extensors are still present in the residual limb (*14*). Commercially available systems that are capable of activating a multiple grips have only recently become available. Classifiers require a rich set of distinct inputs to accurately interpret movement intentions. Existing systems, that use surface EMG, record a spatiotemporal summation of motor unit action potentials from a distant location which is complicated by cross-talk between channels (*15*) and reduced signal strength (*16*).

Furthermore, signals from deep finger flexors (i.e. flexor digitorum profundus to the index finger), are obscured by more superficial muscle activity in the forearm (i.e. flexor digitorum superficialis to the middle finger) or lost entirely in more proximal amputations. In addition to these factors, the reliability of commercial surface electrodes is limited by impedance and position alterations due to sweating or contact shifting (*17*). Hence, distinguishing grasps is challenging for pattern recognition systems that use surface EMG because finger-specific signals are hard to capture and sensitive to environmental changes.

Over the past decade, researchers have investigated software and sensing techniques to improve the reliability of pattern recognition systems and expand their capabilities to include grasp distinction. In earlier work, participants with transradial amputations used surface EMG and pattern recognition to select between 6 and 7 randomly cued hand postures with average movement completion rates of 53.9% (*14*) and 77% (*18*), respectively. Other groups have used alternate machine learning approaches to distinguish up to 10-12 hand and wrist postures with greater than 90% offline accuracy (*19, 20*). The classifiers in these studies accurately distinguished more movements by adding information in the form of multiple input features per channel, or increasing modeling capabilities with deep networks. However, offline simulations alone are not sufficient to characterize online performance as real-time control is heavily influenced by the quality of feedback and individual experience levels (*21*). Pattern recognition algorithms can also struggle to generalize to new contexts. For example, classifiers often issue incorrect predictions while supporting the prosthesis weight in untrained arm positions (*22*). One study found that more robust classifiers can mitigate this issue (*23*). However, this study did not include multiple grasp distinctions. Another study modelled transitions to show that three grasps could be distinguished in a variety of static arm positions and used in a functional task (*24*). Alternatively, another group used sensor fusion of EMG and inertial measurements to control four grasps and hand open. They found that it could improve performance across the workspace (*25*), but required context-specific training. In addition to software innovations, previous studies that quantified multiple grasp control improved signal consistency with adhesive research grade electrodes or captured more information with alternate sensing modalities.

Surgical interventions can be developed to more directly access the peripheral nervous system and improve the quality and fidelity of motor control signals. For users with more proximal amputations (i.e. above elbow amputation), EMG-based systems are limited due to the fact that the muscles used to generate EMG signals for hand control are missing. In these cases, a pattern recognition system would rely on subtle co-activations of remaining musculature, which is inherently difficult. Targeted muscle reinnervation (TMR) is a surgical procedure in which transected peripheral nerves are used to reinnervate surgically denervated areas of muscle. The target muscles are typically superficial and after reinnervation produce functional control signals that can be recorded via surface EMG (*26*). TMR patients with transhumeral amputations or shoulder disarticulations equipped with adhesive electrodes selected four hand postures in a virtual reality environment with an average completion rate of 86.9%. Some patients have been able to control multiple grasps of robotic prostheses in laboratory environments (*27, 28*) and home trials (*13*). In combination with TMR, decomposition algorithms can estimate independent nerve signals from global surface recordings (*15*). This approach requires high-channel electrode grids and real-time control studies have focused on separation of wrist and gross hand movements to date (*29*). Implantable electrodes have also been proposed to record efferent motor commands directly from individual nerves. However, these electrodes are limited by low amplitude efferent motor action potentials (*30*) or lack of chronic stability. Conversely, electrodes implanted into muscle tissue can record strong and stable EMG signals. In one study, indwelling electrodes eliminated the need to re-calibrate a regression controller in a virtual reality environment for months (*31*).

Intramuscular electrodes have also been shown to improve signal strength and control precision independent of changes to the mechanical interface or control strategy (*16*). In another study, electrodes with multiple contacts were implanted into individual muscle bellies, enabling 6 degree of freedom (DOF) dexterous control including individual fingers of a robotic hand (*32*). However, the ability for intramuscular electrodes to capture finger movements requires a sufficient amount of residual innervated musculature or other surgical interventions.

At the University of Michigan, we have developed the Regenerative Peripheral Nerve Interface (RPNI) which consists of a free muscle graft sutured to the end of a divided peripheral nerve. The muscle graft regenerates and becomes reinnervated by the regenerating peripheral nerve axons. RPNIs have been shown to effectively prevent and treat neuroma pain and phantom pain (*33, 34*). Most importantly, RPNIs serve as a stable biological amplifier for efferent motor signals, retaining anatomical consistency between the host nerve and RPNI functions (*35*–*37*). In a clinical trial, high resolution control signals recorded from electrodes implanted into RPNIs remained stable for up to 300 days (the duration of observation)(*38*). Vu et al. also used a pattern recognition algorithm in a virtual environment to validate RPNI control capabilities (*38*). Pattern recognition is the state of the art for intuitive device control and a natural match for most multi-articulating hands that are designed to switch between grip patterns. Therefore, it is both an easily comparable and immediately applicable control paradigm for implantable technologies.

In this study, we continue experiments with two persons with transradial amputations and electrodes surgically implanted into RPNIs and residual innervated muscles. Vu et al. focused specifically on each participant’s RPNIs, here we investigate the signal quality and control capabilities of the full set of RPNIs and residual innervated muscles. We demonstrate that indwelling electrodes captured stable EMG control signals from both RPNIs and residual muscles with a highly favorable Signal-to-Noise ratio (SNR) in both participants. In addition to large amplitude recordings, the EMG was also highly specific for motor decoding. Real-time control with classifiers often presents a trade-off between responsiveness and stability when determining the length of the EMG processing window (*39*). Here we find a more optimal solution to this dilemma and utilize a Hidden Markov Model (HMM-NB) which learns transitions between latent states to make rapid predictions without sacrificing temporal stability (*40*). We demonstrate online performance in a virtual posture switching task designed to reflect real-time finger and grasp control. To our knowledge, this is the fastest and most accurate signal acquisition and pattern recognition system that can switch directly between individual finger movements. We quantified performance for two posture sets: a 10 class set including extrinsic and intrinsic individual finger and hand movements and wrist flexion, and a five class set of functional grasps. To demonstrate robustness across physical contexts, we quantified stability of a grasp classifier in one participant across novel static arm postures with a prosthesis donned. Finally, both participants completed a functional assessment where they used robotic prostheses to move differently shaped objects that required the use of multiple hand grasps.

## Results

### Speed and Accuracy of Finger and Grasp Classifications

Two persons with transradial amputations (P3 and P4 from Vu et al., labelled P1 and P2 here), underwent surgery to have eight pairs of bipolar electrodes (Synapse Biomedical, Oberlin OH) implanted into RPNIs and residual innervated muscles. Differentiated signals from each electrode pair were filtered and the mean absolute value (MAV) was extracted with a sliding window. A Hidden Markov Model (HMM-NB) was trained by instructing participants to mimic 5-7 repetitions of each posture with their phantom limb. In offline simulations, the HMM-NB distinguished the same individual finger and wrist movements, known as the “1 of 10” posture set in P1 and P2 with 95.4% and 94.1% accuracy (Fig. 1A,D). Additionally, both participants controlled a virtual hand in real-time with the HMM-NB to complete a fast-paced posture switching task between each of the 10 postures. In one experiment session, P1 successfully maintained a one second hold within the 10 second timeout period on 100% of his trials. Online classifier accuracy measured real-time control errors during the first second of transition to the cued postures. P1 achieved an online accuracy of 93.0% in his experiment session (Fig. 1B).

**Fig. 1.**
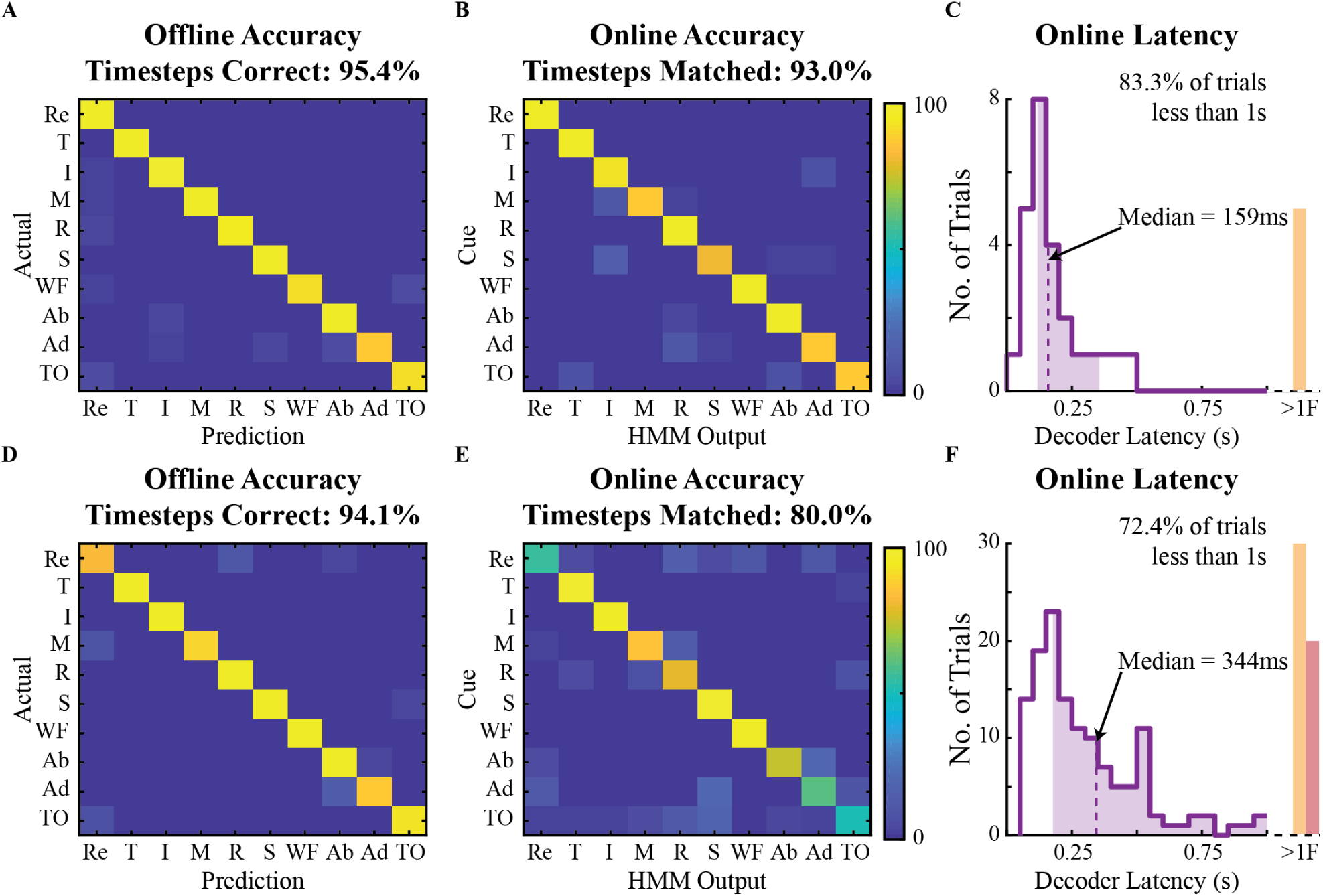
Decoding individual finger and wrist postures. Participants controlled a virtual hand with the Hidden Markov Model (HMM-NB) to match 10 postures: flexion of all five fingers (T,I,M,R,S), wrist flexion (WF), finger abduction (Ab), finger adduction (Ad), thumb opposition (TO), and rest (Re). **(A)** Simulated offline performance of the HMM-NB during rest and hold periods for P1’s training data (5-fold cross validation, 5-6 repetitions per movement). **(B)** An online confusion matrix captures transition errors to cued postures while P1 controlled the virtual hand in real-time. **(C)** Decoder latency was measured as the time difference between the onset of new EMG activity and a successful posture. The median (dashed line) and middle 50% (shading) is overlaid on histograms binned in 50 ms increments (n = 30 trials). Trials with latency greater than a second (>1) are aggregated in the orange rectangle. **(D)** Offline confusion matrix for P2 using training data from one experiment session. **(E-F)** Same as above for P2 across three single day experiment sessions (n = 181 trials). Trials where a one second continuous hold could not be maintained are marked as failures (F) and aggregated in the red rectangle.

As shown in Fig. 1C, P1 used the HMM-NB to rapidly switch between postures with an online latency of 159±237 ms (median±i.q.r.). P2 performed the same calibration and decoding routine for three single day experiment sessions for the 1 of 10 posture set. Across all sessions, she achieved an online accuracy of 80.0% with a latency of 344±924 ms (Fig. 1D,F). P2 was able to make corrections and maintain successful holds on 89.3% of her trials. She had the most difficulty performing finger adduction, thumb opposition, and relaxing to rest with the virtual hand.

Unsurprisingly, online performance improved with a reduced number of postures (Fig. 2). Both P1 and P2 completed the same set of experiments using the HMM-NB to rapidly switch between five postures: three functional grasps, finger abduction, and rest. Offline accuracy was relatively low for P2, notably the HMM-NB issued incorrect predictions of rest during simulated holds. However, when controlling the virtual hand in real-time, P1 and P2 achieved online accuracies of 99.5% and 96.3% across one and three experiment sessions respectively. Both participants were able to recover from errors and maintain successful holds on 100% of their trials. The HMM-NB was also able to distinguish the grasps posture set with even lower median latencies of 96±30 ms and 173±151 ms (median±i.q.r.) for P1 and P2 respectively. To compare to previous studies that classified finger and hand postures, movement completion metrics were calculated ignoring wrist flexion and rest trials (Table S1).

**Fig. 2.**
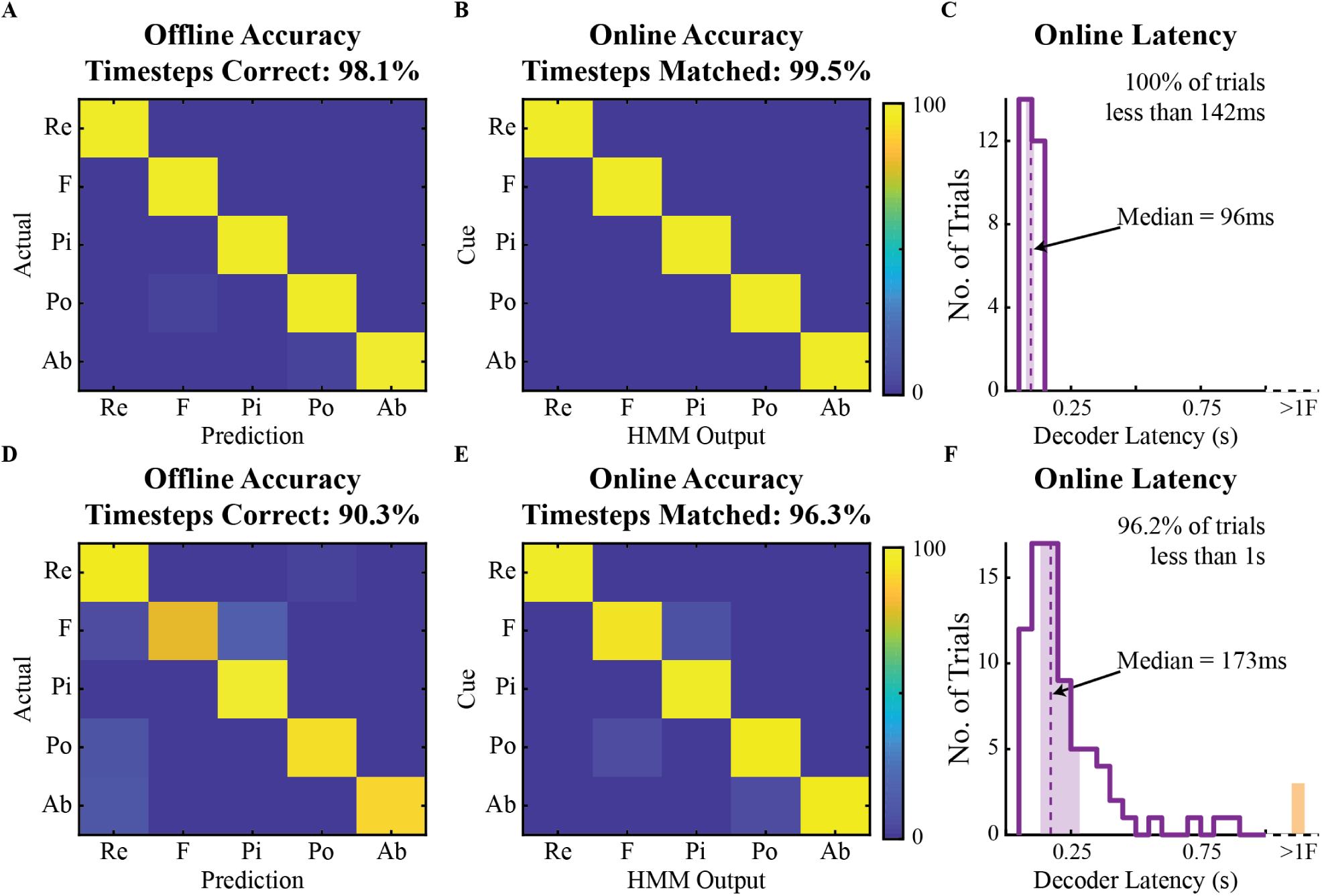
Decoding hand grasps. A fewer number of functional grasps could be predicted in real-time with higher online accuracy and lower latency. **(A)** Simulated offline performance of the HMM-NB output distinguishing fist (F), pinch (Pi), point (Po), finger abduction (Ab), and rest (Re) for P1 (5-fold cross validation, seven repetitions per movement). **(B-C)** Online accuracy and decoding latency for P1 measured as in Fig. 1 (n = 26 trials). **(D)** Offline confusion matrix for P2 using training data from one experiment session (5-fold cross validation, five trials per movement). **(E-F)** Same as above for P2 across three single day experiment sessions (n = 79 trials). Trials with latency greater than a second (>1) are aggregated in the orange rectangle.

### Robustness Across Arm Positions

In order to be of practical use for patients with upper limb amputations, decoders must be reliable to different physical contexts. P2 donned an i-Limb Quantum™XS (Ossur, Reykjavik, Iceland) and was able to switch between functional grasps in eight different arm orientations. P2 had limited elbow range of motion, described in Supplementary Materials and Methods, and was asked to match arm postures to the best of her ability. The virtual reality environment was again used for cues and visual feedback to quantify decoding performance across 20-22 trials per arm position (Fig. 3A). The HMM-NB used for this exercise was trained with her arm and prosthesis resting on the table. Classifier performance remained robust across the majority of the arm positions where transition errors were infrequent and quickly corrected (Fig. 3B,C). In six of the eight arm positions, P2 was able to control the virtual hand with online accuracy never dropping below 96% and a decoding latency of 173±113 ms (median±i.q.r). In two arm positions, arm raised and behind the back, there was a noticeable increase in transition errors to the cued posture, which occurred 7 and 6 times respectively. These positions at the superior and posterior extremes of her workspace had lower classification accuracy and a higher decoding latency of 301±299 ms. Even though there were more frequent errors in these two positions, the classifier did not completely fail and P2 was able to recover and achieve a successful one second hold within the timeout period on 100% of trials. In the same session. P2 used the same decoder to activate grips with her robotic prostheses following verbal instruction in five of the arm positions. During preliminary testing and demonstrations, P1 was able to activate the same set of postures using the LUKE arm (Mobius Bionics, Manchester NH) in three arm positions. P2 also activated individual flexion of fingers on the i-Limb in three arm positions, correctly executing 14 out of 15 flexion attempts.

**Fig. 3.**
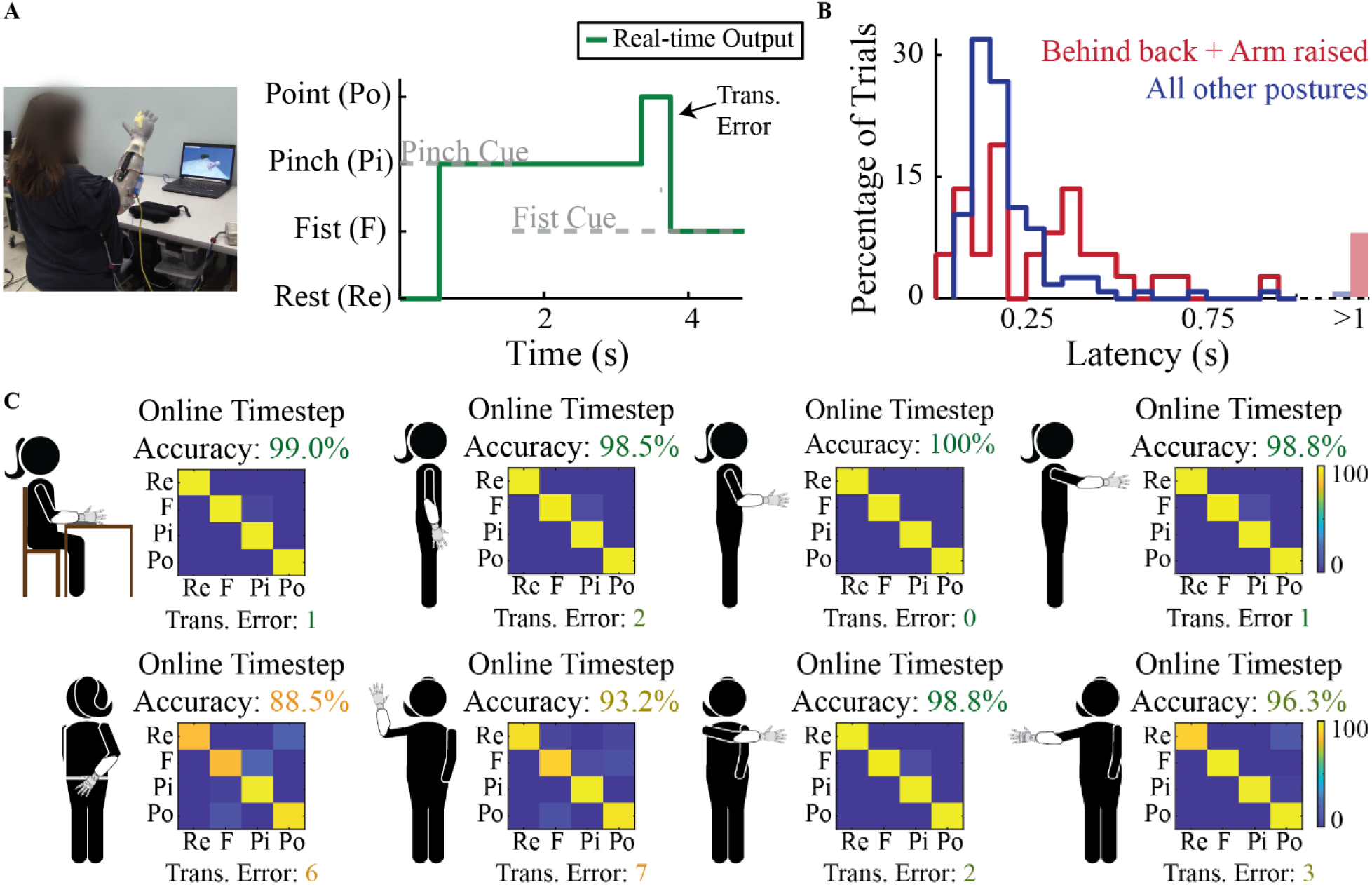
Robustness across static arm positions. P2 performed control experiments in the virtual reality environment to quantify decoder performance across eight different arm positions. **(A)** P2 donned her prostheses, but used the virtual reality environment to switch between fist (F), pinch (Pi), point (Po) and rest (Re). **(B)** P2 calibrated the HMM-NB with her arm resting on a table. Latency histograms binned in 50 ms increments for the majority of positions aggregated (blue, n = 116 trials), as well as the two positions with more frequent and longer transition errors (red, n = 37 trials). **(C)** In addition to online accuracy, the number of transition errors was also reported for each of the 8 positions.

### Functional Prosthesis Use

Both participants used the functional posture set to complete five trials of a Southampton Hand Assessment Procedure (SHAP)-inspired task (Fig. 4). Each trial consisted of five object interactions and required the activation of three different grasps. Participants were instructed to use specific grasps for the timer and each SHAP object. P1 used the LUKE arm to move SHAP heavy objects and completed the task with an average time of 18.75±3.42s (mean±s.t.d.). P2 used the i-Limb Quantum™to move SHAP light objects and completed the task in 36.60±7.66s on average. P2 completed the objects in the reverse order. This made it easier for experimenters to assist with manual wrist adjustment for the power object. Both participants were largely successful, performing 23 out of 25 interactions without dropping an object or failing to press the button.

**Fig. 4.**
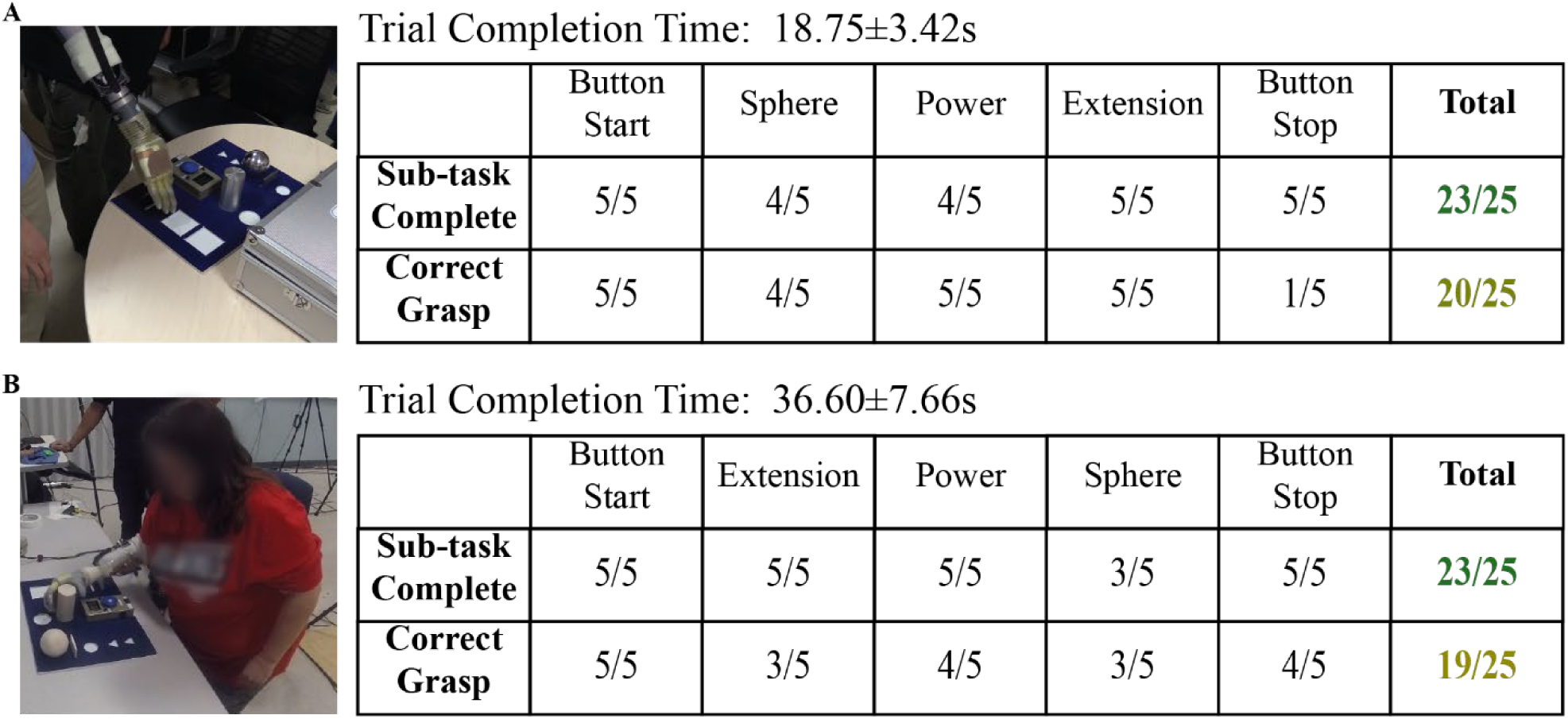
SHAP inspired grip switching task. Both participants used their prostheses with the functional posture set to perform a task in which they were instructed to use specific grasps to interact with three objects and a timer. Sub-tasks were counted as complete if the participant succeeded on their first attempt, and correct grasps were counted when the instructed grasp was achieved on the first EMG activation attempt. **(A)** P1 used the LUKE arm and was instructed to use point to start the timer, fist for sphere, fist for power, and pinch for extension objects, before using point to stop the timer. **(B)** P2 used an i-Limb Quantum XS and completed the object trio in the reverse order. Completion time (mean±s.t.d, n = 5 trials) included manual wrist adjustments for P2.

Additionally, P1 and P2 performed the instructed grasp during their first EMG activation attempt on 20 and 19 out of 25 interactions. The main issue P1 encountered was an inability to use point for timer stop. Interestingly, this error never occurred at trial start indicating the point could be activated under the right conditions. Both participants had instances where they led with a pinch to pick up the sphere. P1 seamlessly corrected and engaged the instructed fist grasp in all but 1 trial, while P2 needed to perform additional EMG activations for correction. The i-Limb took longer to switch between grips, so a state machine was coded that required P2 to issue a rest command before activating a grasp. This and the need for manual wrist adjustment contributed to the variation in completion times between participants.

### Signal Strength and Specificity

The bipolar intramuscular electrodes recorded specific and spatially segregated EMG activity particular to individual finger movements. To visualize posture segregation, we used linear discriminant analysis (LDA) to define the top two discriminating dimensions for the 1 of 10 posture set (Fig 5). For this analysis, MAV from the 8 bipolar electrode pairs were processed with time history parameters that were input to a standard Naive Bayes classifier for successful real-time control (Table S1). Rest data was excluded to maintain focus on movement distinctions. Overall, posture holds were well separated in the low dimensional space on both real-time and trial-averaged timescales. For P1, thumb movements were the most distinguished from other postures and separated along the first discriminant axis. For P2, wrist flexion was the best separated posture, along with flexion of the thumb or index finger in the first two discriminant dimensions. These movements were amongst those explicitly targeted by electrode implantation and were well represented by individual channels (Fig. S3 and Fig. S4). Additional discriminating dimensions would further separate movements, with 4-D embedding capturing approximately 90% of the variance in training data.

**Fig. 5.**
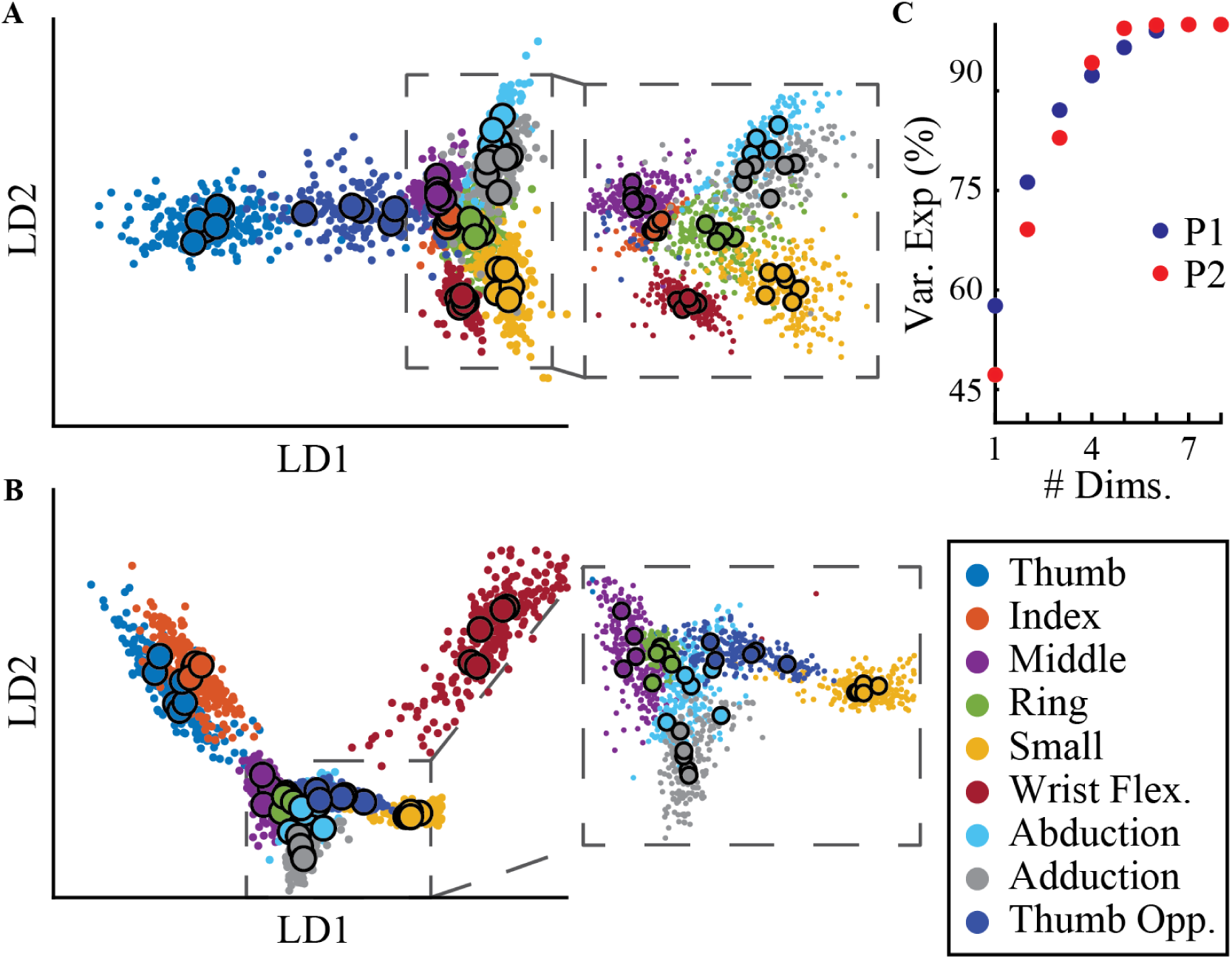
Dimensionality reduction for posture visualization. MAV from eight channels reduced to two discriminant dimensions found using LDA. Large outlined points indicate trial averages, while small dots represent individual timesteps during posture holds. Dashed boxes magnify separation of relatively close movements. (A)MAV for P1 was processed in non-overlapping 50 ms time bins. **(B)** MAV for P2 was processed in a sliding 200 ms window updated every 50ms. **(C)** Cumulative variance explained by additional dimensions.

In addition to high specificity, the implanted electrodes also recorded large amplitude responses with a low noise floor (Fig. 6). For P2, we compared the SNR of five intramuscular channels to simultaneously recorded surface signals. Bipolar surface recordings were acquired using adhesive gelled electrodes (Biopac, Goleta, CA) connected to the same signal processing equipment. Across the compared channels, the average SNR for implanted electrodes was 105.4±82.6 and 152.5±138.6 (mean±s.t.d) gain for P1 and P2. P1 did not participate in a surface session, while P2’s surface signals averaged an SNR of 8.5±6.6. A high surface SNR of 19.6 was observed during finger abduction which prompted a full splay of the hand. We recorded this movement with surface electrodes targeting extensor pollicis longus (EPL) as described in Supplementary Materials and Methods. However, it is possible we also recorded activity from extensor digitorum communis (EDC) or other nearby muscles. Deep muscles that control individual finger movements were more difficult to capture from the surface. Gelled electrodes targeting flexor digitorum profundus of the index finger (FDPI) recorded signals with an SNR of 2.26 during index finger flexion. The SNR for simultaneously recorded implanted electrodes was 244 for EPL and 66 for FDPI. We also measured SNR for electrodes implanted into RPNIs, however attempts to accurately target and record matching surface EMG were unsuccessful. By nature, RPNIs also do not have a direct functional correspondent, so residual muscles that control related movements were selected for comparison. Across all the comparisons conducted with P2, implanted electrodes provided a 6 to 30-fold SNR improvement.

**Fig. 6.**
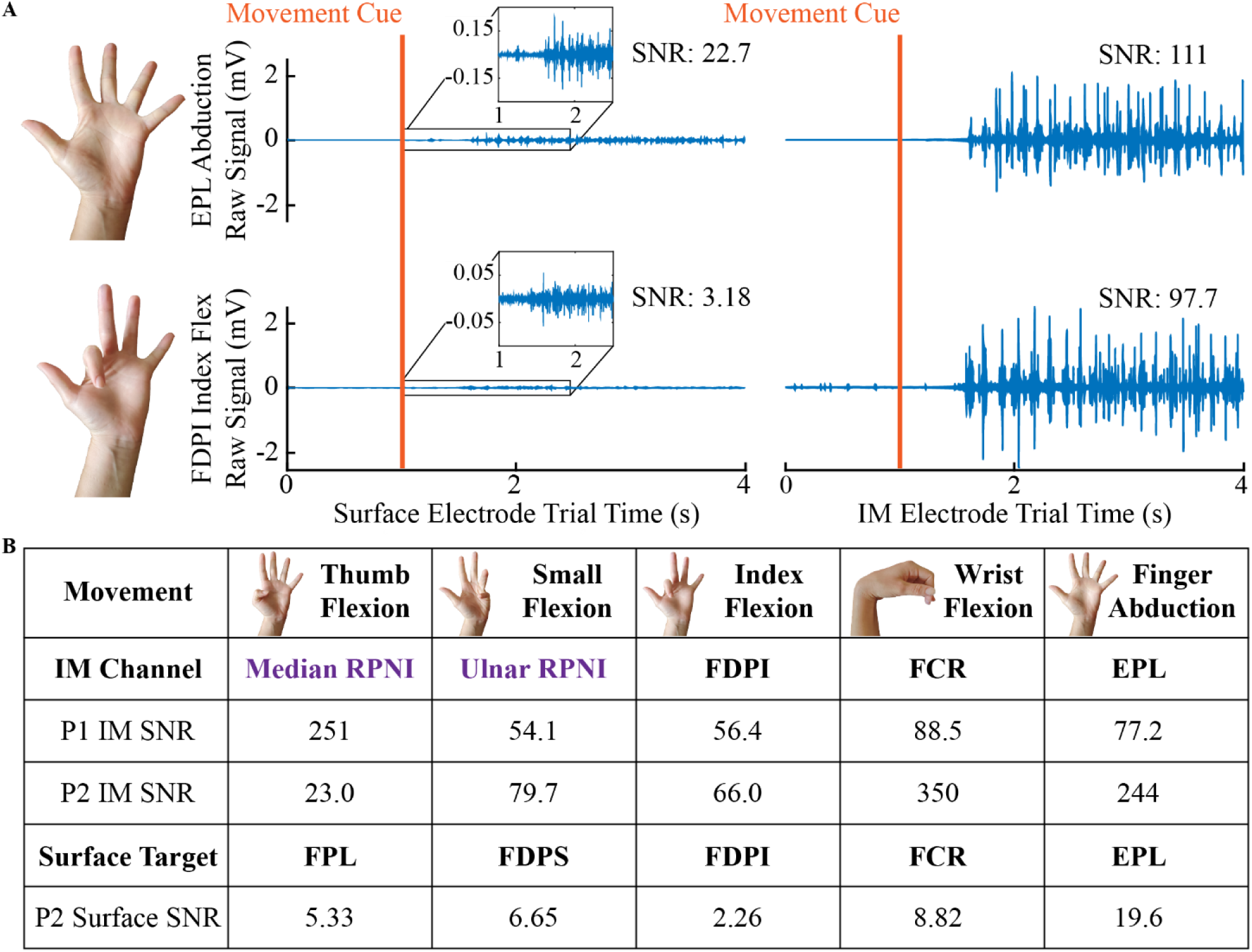
SNR of intramuscular and surface EMG for finger movements. Signal-to-Noise Ratios (SNR) from implanted (IM) electrodes from both subjects and simultaneous surface recordings from P2. **(A)** Example SNRs from individual movement trials with simultaneously recorded surface and implanted electrodes from P2. For both movements, the trial with the highest surface SNR was chosen. (B)SNRs were calculated for implanted channels by averaging the active and rest RMS voltages across 5 trials of the individual finger movement most relevant to the targeted muscle. For P2, simultaneous surface recordings were obtained by individually targeting implanted residual muscles. For RPNI comparisons (purple), appropriate residual muscles were selected based on the cued movement.

### Alternate Classifier Simulations

The HMM-NB models underlying state transitions to rapidly issue accurate predictions. Classification stability and accuracy can also be improved by adding time history and additional informative features. Fig. 7 demonstrates the offline ability of the HMM-NB and a standard Naïve Bayes (NB) classifier as well as three alternate classifiers distinguishing the 1 of 10 posture set. The alternate classifiers used 5-time domain features and 6^th^ order auto-regressive coefficients to characterize EMG from each channel. Increasing window length did not create multiple time points for input, preventing alternate classifiers from modelling EMG patterns over time. P1’s alternate classifiers yielded improvements in combination with larger processing windows, however this difference was not robust for P2 across four training sessions. Overall, the performance difference between NB using only MAV and each alternate classifier was not significant for implanted electrodes (p > 0.2, paired t-test, n = 35 window lengths across 5 datasets). However, additional features proved beneficial when applied to P2’s surface EMG, which NB could not well distinguish from only MAV. Adding features increased P2’s surface performance by 17.8±10% (mean±s.t.d.) across all window lengths without changing the classification model. By comparison, additional features did not significantly impact NB performance with the implanted electrodes (p > 0.5, paired t-test, n = 35 window lengths across 5 datasets). The HMM-NB consistently improved simulated performance over NB, and overall outperformed each alternate classifier (p < 0.01, paired t-test, n = 42 window lengths across 6 datasets). The HMM-NB most noticeably improved performance for smaller processing windows. These results indicate that algorithms modelling state transitions or EMG dynamics can improve accuracy and remain robust to smaller processing windows. This potentially benefits real-time control applications by reducing a trade-off between responsiveness and stability. An online comparison between the NB classifier and the HMM-NB is detailed in Supplementary Materials and Methods. Both participants used the NB classifier to complete the 1 of 10 posture switching task with average online accuracy of 85.8% and latency of 328 ms. These results are promising for situations that demand simplicity or are highly sensitive to computational workload.

**Fig. 7.**
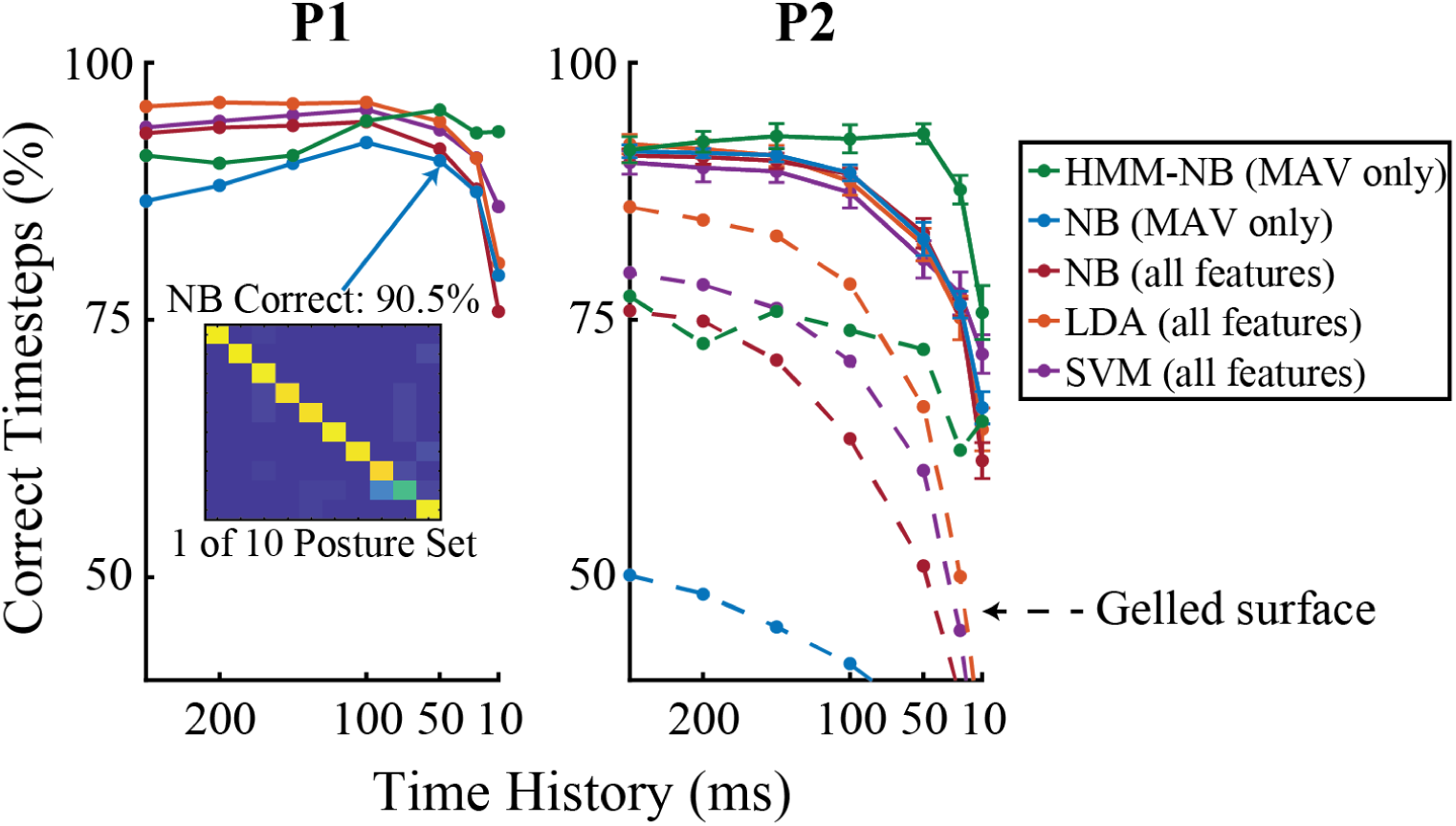
Offline classifier simulations. Offline performance of different algorithms evaluated across 7 increasing window lengths on the 1 of 10 posture set for P1 and P2. The Hidden Markov Model (HMM-NB) and a standard Naïve Bayes classifier (NB), used only mean absolute value (MAV) as an input feature. An alternate NB implementation, linear discriminant analysis (LDA), and a support vector machine (SVM) used five time domain features and 6^th^ order auto-regressive coefficients per channel. Classifiers were evaluated across individual timesteps during hold and rest periods using 5-fold cross validation. Dashed lines for P2 represent performance using EMG recorded from eight bipolar gelled adhesive surface electrodes. Performance with P2’s implanted electrodes (mean±s.e.m) was evaluated for simultaneously recorded intramuscular EMG and three calibration sessions used for real-time control experiments.

## Discussion

In this study, we have demonstrated that implanted electrodes record high-quality EMG signals from RPNIs and residual innervated forearm muscles in two persons with transradial amputations. Both participants were implanted with electrodes targeting the same individual finger movements and were able to control a virtual hand to distinguish the same posture sets which included individual finger, intrinsic, and compound hand movements. The posture switching task tested real-time control capabilities of the high speed pattern recognition system, allowing users to directly switch between postures and rest. The speed and accuracy of our pattern recognition system with implanted electrodes, to distinguish finger movements, exceeds earlier work which quantified real-time performance in virtual environments (Table S3). In a controlled environment, the HMM-NB also distinguished a smaller set of functional postures in novel static arm positions.

P2’s performance was consistent across the majority of the workspace, which we attribute to the stability of signals from the implanted electrodes. Finally, we demonstrated that participants could use the high speed pattern recognition system to control advanced robotic prostheses, eliminating the need for time consuming grip triggers or selection schemes. The LUKE arm came equipped with real-time research software that allowed P1 to achieve functional control similar to the virtual environment. He was able to activate grasps without a perceived delay and seamlessly switch between postures. P1 leveraged this capability to use the pinch and fist grasps to manipulate the SHAP sphere. For P2, our pattern recognition system was adapted to work with commercially available grip selection software, similar to previous work (*24, 25, 27*).

Enforcing a grip entry/exit structure can improve stability, however it increases the effort required to correct erroneous outputs. Furthermore, the ability to move directly between hand postures has been shown to improve functional performance (*28*). Regardless of the hand interface, implementing proportional control will be beneficial for more precise tasks. The specificity of the indwelling electrodes offers the ability to directly map muscle activations to individual fingers. Different mappings can be explored to either improve precision within i-Limb grips or increase dexterity of the LUKE arm during object manipulations.

Pattern recognition is just one approach being considered for intuitive grasp control, although it may be the most practical and immediately applicable technique, because it pairs well with commercially available hand software. Furthermore, classifiers can be computationally inexpensive and remain accurate when increasing the number of predicted movements relative to the number of input channels (*14, 24, 25*). For example, in Vu et al., P1 was able to activate rest and four finger postures using only two EMG channels (*38*). However, it can be challenging for classifiers to generalize to new contexts. In both participants, arm movements occasionally produced unintentional extensor activity. Ignoring unreliable channels was a quick and effective solution for this study, but could limit the number of predictable grasps compared to more robust classifiers (*23*). Changing EMG patterns during object interactions also led to some misclassifications when P1 pressed the timer button with the point grasp. Similar phenomena have been noted in other research applications (*41, 42*) and could be a natural product of the motor system (*21, 43, 44*). Regression algorithms are being explored in combination with implantable electrodes for multiple DOF hand control (*31, 32, 38*). In addition to increased dexterity, regressors may be more robust to changing contexts (*45*). However, there are limitations on the number of DOF that can be simultaneously and independently activated. Biomechanical control strategies are also being developed that use a principled musculoskeletal model to provide robust estimates of intended joint kinematics and torques (*29, 46*). In order to be effective, regression and biomechanical techniques require the prosthetic hand to allow precise and simultaneous control of individual fingers. The ability to incorporate an active wrist is also an important consideration for algorithm development, as it allows for more natural body movements (*47*). Ultimately, the signal quality and strength of implanted electrodes is relevant to any EMG control paradigm. Our signal processing and feature extraction provide a stable proxy for local motor unit activity for residual muscles or the peripheral nervous system via RPNIs (*36, 38*). Signals from implanted electrodes can be stable for months to years (*31, 38*), which is advantageous for pattern recognition and regression algorithms as large calibration datasets can readily be assembled.

Our experimental system used indwelling electrodes that exited the skin and were connected to a computer. While we have found these signals stable over years, this is not a viable long-term solution for most users. The signal processing chain can readily be adapted to a low-power architecture (*48*), however many other system features need to be considered. Multi-channel percutaneous electrical connectors have been developed (*49*), but risks of complication and infection have limited their use to temporary or life-saving medical devices (*50*). Osseointegration is a hardware innovation that improves the mechanical linkage between user and prosthesis. In our study, we observed unintentional muscle activations during some arm movements and reduced performance at the extremes of P2’s range of motion. For some patients, osseointegration may mitigate these issues and improve controller reliability by reducing the effects of prosthesis weight. It also has the benefit of creating a direct physical feed-through for intramuscular electrodes (*16, 51*). However, it is limited to patients that meet anatomical criteria such as residual limb length. Fully implantable wireless devices have gained traction for neuromodulation therapies (*52, 53*). By modifying these approved neuromodulation systems, development and certification costs can be reduced by utilizing shared platforms across the industry (*54*).

This study focused on expanding the capabilities of pattern recognition algorithms to extract hand motor commands reflective of user intent. However, a lack of sensory feedback and proprioception can limit the benefits of myoelectric devices. For example, even though thumb opposition and finger abduction were accurately predicted offline, P2 had difficulty activating these postures during real-time control. P2 reported limitations on proprioceptive feedback in her phantom limb, notably including her thumb metacarpal phalangeal joint. Additional practice (*21*) or enhanced training strategies (*55*) could potentially resolve this issue. However, improved feedback mechanisms (*56*–*59*) may be necessary for some patients to realize the full potential of improved motor control.

Increasing the responsiveness, precision, and sensory feedback can provide additional benefits for prosthetic users such as increased trust (*16*), increased embodiment or reduced phantom limb pain in some users (*60*). Regardless of the specific algorithm or control strategy, the stable and highly specific EMG afforded by indwelling electrodes can play a significant role in improving prosthetic control and user satisfaction in the coming years.

## Materials and Methods

### Signal Processing and Experiment Set-Up

The two participants with RPNIs and indwelling electrodes labelled P1 and P2 for this study, were respectively labelled P3 and P4 in Vu et al. (*38*). Detailed anatomical information is also present in Supplementary Materials and Methods. Briefly, both participants had bipolar electrode pairs (Synapse Biomedical, Oberlin, OH) chronically implanted into their ulnar and median RPNIs along with the following residual muscles targeting finger and wrist functions: Flexor Pollicis Longus (FPL), Flexor Digitorum Profundus - Index Finger (FDPI), Extensor Pollicis Longus (EPL), Extensor Digitorum Communis (EDC), and Flexor Carpi Radialis (FCR). P1 also had an electrode pair implanted in Flexor Digitorum Profundus - Small Finger (FDPS). EMG from the implanted electrode contacts was amplified, sampled at 30kSps, and referenced with a Cerebus Neural Signal Processor (Blackrock Microsystems, Salt Lake City, UT). The Cerebus simultaneously recorded and sent referenced EMG to a Matlab xPC (Mathworks, Natick, MA) for real-time processing. Fig. 8D shows the signal processing chain on the xPC for online decoding. The xPC applied the 100-500Hz band-pass filter and down-sampled EMG to 1 kSps, from which mean absolute value (MAV) was the only feature extracted for online decoding. The xPC sent UDP packets to manipulate two virtual hands (*61*) which were presented to the subject on an external laptop. A lead hand to cue postures was positioned in the foreground, while a secondary hand which the subjects could control was positioned in the background. The xPC’s real-time guarantee ensured that EMG was processed and time-synced with behavioral data and decoders executed within one millisecond.

**Fig. 8.**
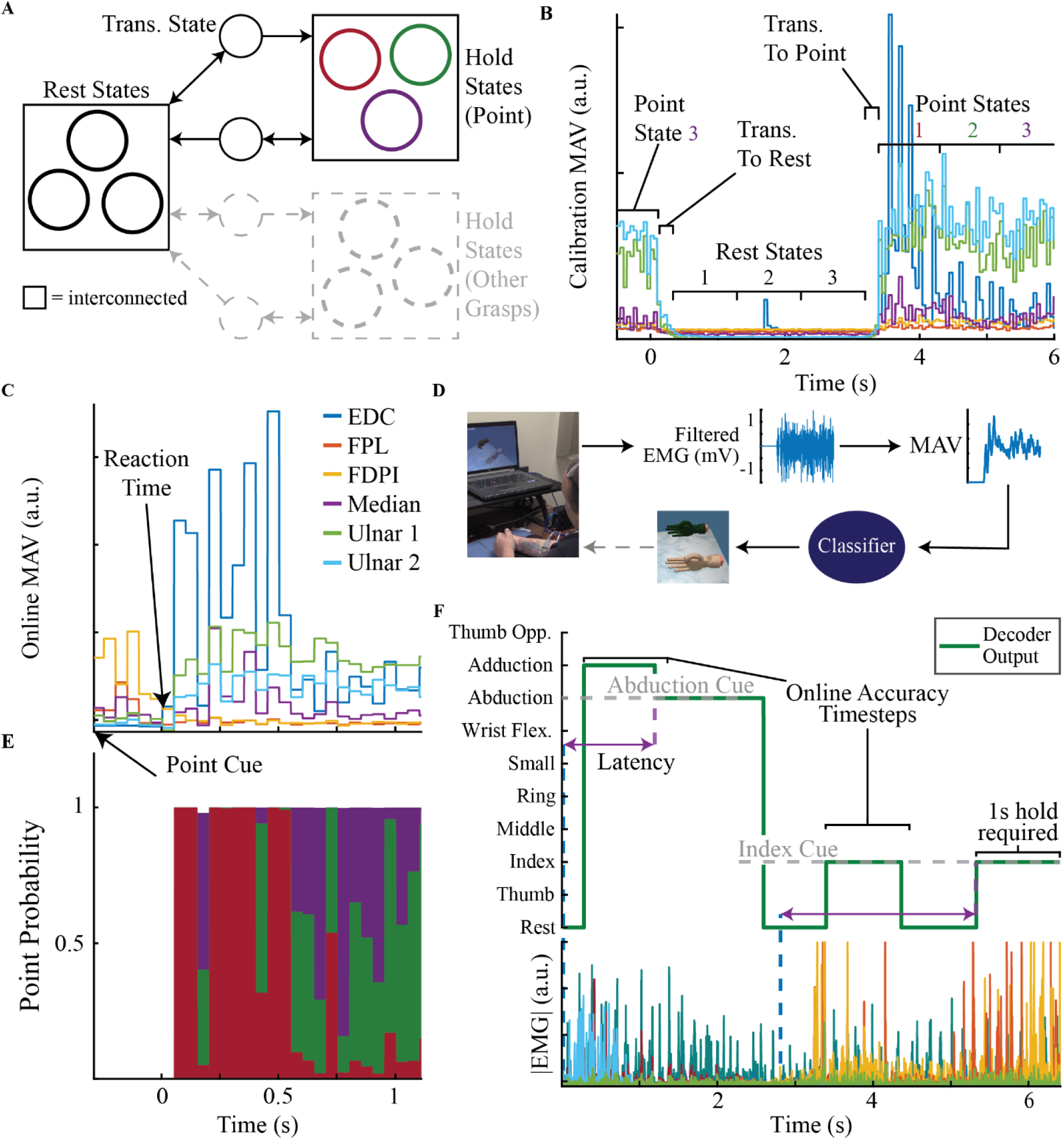
Posture switching task and performance metrics. **(A)** The HMM-NB contained three interconnected states for rest and posture holds with one transition state to and from rest. Point highlighted for example. The probability of a posture was determined by summing the probabilities of the rest and hold states. **(B)** An expectation-maximization unsupervised learner was carefully initialized by placing rest and hold states sequentially along active hold periods with transition states in between. Mean absolute value (MAV) was the EMG feature used for online decoding. **(C)** P2’s online activity during a switch to point posture. **(D)** Participants controlled a secondary hand (background) with visual feedback to match posture cues (foreground). **(E)** Optimized latent states of the online decoder capture EMG dynamics in C. One state (red) reflects a sharp increase in EDC activity while two states (green and purple) capture continued hold of the Ulnar RPNIs. **(F)** Participants were required to hold the cued posture for one second continuously before the next cue was presented. Decoder latency measured classifier responsiveness as the time difference between new EMG activity and a successful hold. Online accuracy was evaluated across individual timesteps for the first second of transition to the cued posture. 1 of 10 posture set with errors shown for example.

### Hidden Markov Model for Real-Time Control

Many EMG classification algorithms are error prone when users move between postures or during extended holds. This is because they characterize postures as an average point in feature space and have no knowledge of the prior state or capability to model transitions. Common methods to improve performance are to increase the length of the processing window for stability or add additional output filters to reduce transition errors. These methods are effective, but they automatically introduce a compromise between classifier stability and responsiveness (*39*). Here, we explore a more principled solution. The Hidden Markov Model (HMM-NB) models transitions between latent states and closely resembled previous work (*40*). The probability of a latent state occurring is determined both by the observed EMG inputs as well as the occurrence of previous states. A linear transition matrix determined the likelihood of moving from one latent state to another and optimally smoothed transitions. A Naive Bayes model represented three interconnected “hold” states per posture which can capture dynamic EMG patterns. The model also contained “transition” states going to and from rest as shown in Fig. 8A. State models were carefully initialized by splitting the active period or rest periods of the training data into thirds Fig. 8B. Active periods were automatically marked in training data per trial by retroactively finding active channels and selecting the starting point of when their MAV exceeded 4 times the standard deviation of the trial’s rest data for at least 200ms. The transition matrix for “hold” and “rest” states was initialized with an equal probability of remaining in or moving to another connected state. “Transition” states were initialized with a 0.9 probability of remaining in the state. We limited the expectation-maximization algorithm (*62*) to five iterations and selected parameters from the iteration with the highest likelihood on held out test data. During online decoding, the probability of a given posture or rest was taken as the sum of the three “hold” or “rest” state probabilities as shown in Fig. 8E. The “transition” states were not selected for output because they represented the beginning of activations from rest, while participants could directly switch between grasps during real-time control. Fig. 8C shows an example of P2 initiating a point during online control. The solution found by the HMM-NB captures her changing EMG patterns, as shown in Fig 8E. The inclusion of multiple latent states per posture increases modeling capabilities and the ability to model transitions allow the HMM-NB to operate with less time history than steady-state classifiers. Performance comparison to a standard NB classifier and analysis are included in Supplementary Materials and Methods.

### Virtual Posture Switching Task

Table S2 shows the posture sets explored in this study. MAV from all eight channels and calculated with 50 ms of time history, was used to control the virtual hand for both the 1 of 10 and grasps posture sets. P1 had significant time constraints during the course of the study. He performed one session of the virtual posture switching task for both the 1 of 10 and grasps posture sets. P2 completed three sessions for both posture sets. To calibrate the HMM-NB, participants matched the virtual cue hand with their phantom hand to the best of their ability. Participants faced the external laptop while resting their arm on the table in a neutral and comfortable position. Calibration runs involved participants holding each of the desired postures 5-7 times for 2.5-3 seconds with an equal length rest period in between each hold. Where possible, calibration runs were completed on the same day before online control runs. Even though the total routine was relatively quick (approximately five minutes or less), older calibrations – 18 days for P1’s 1of 10 posture switching task and 16 days for P2’s arm position test – were used twice to save time and effort. During online real-time control, participants actively controlled the secondary hand and attempted to match the cue hand. Participants used a free running classifier, meaning the decoder output was never automatically reset to rest or a particular posture. The secondary hand turned green when the decoder output matched the cued posture as an additional success indicator. Participants were required to match the cued posture for one second without interruption. If the participants failed to achieve a one second hold within 10 seconds, the trail was considered a failure. A five second timeout was used for P2’s arm position test. After a successful hold or failure, a new posture was immediately cued in a pseudo-random order. This task encourages participants to directly switch between postures at a faster pace than the training procedure. The requirement of a continuous hold ensures that successful algorithms must be both stable and responsive. In summary, the posture switching task was designed to be indicative of a classifier’s capability to actively control prostheses in real-time.

Fig. 8F highlights two of P2’s real-time control trials with errors to demonstrate the analysis metrics for the posture switching task. Online classification accuracy was evaluated across individual timesteps for the first second of transitions to the cued posture. Perfect accuracy required the classifier to switch and maintain a one second successful hold without any error. The one second hold length was chosen to be similar to the selection period in previous work (*14, 27*). Sometimes, participants would pause before attempting the cued posture, possibly due to physical or mental fatigue. Rest outputs under those circumstances were ignored, however moving from the cue to rest was penalized. Decoder latency measured classifier responsiveness and controllability.

Latency was calculated as the time difference between the onset of new muscle activity and the beginning of a successful hold. EMG onset was determined visually by viewing the filtered and rectified EMG for each trial and marking the beginning of a new EMG pattern. EMG onset approximates reaction time and averaged 604.9±268.8 ms for P1 and 517.8±281.6 ms for P2 (mean±s.t.d.). Latency was not calculated for trials without a distinct EMG change or when the pseudo-random order produced a duplicate. These were rare occurrences, accounting for 3.5% of all trials. Failed trials were marked with a 10 second latency to reflect task timeout. Median and i.q.r. were used to characterize latency since the distributions did not have a normal shape.

### Functional Testing

The functional posture set was used to test decoder stability across arm positions and to complete the SHAP inspired task. P2 completed one arm position session quantified with virtual posture switching task described above. Both participants performed the SHAP-inspired task in one session. The functional posture set matched the direct control strategy for robotic prostheses which used rest to open the hand. We did not employ a distinct hand open state because the robotic hand interface was sensitive to instabilities. We observed instances of unintentional extensor activity during arm movements, which could erroneously trigger finger abduction predictions. Incorporating an active hand open signal for proportional control is a consideration for future studies. P2’s functional decoder used MAV from her RPNIs, FPL, FDPI, and EDC. EPL was observed to activate unintentionally during arm movements and held out. FCR was not observed to cause any issues but was removed as a precaution since it was not necessary to distinguish grasps. P1 used a decoder that received MAV only from his RPNIs, FPL, FDPI, and FDPS electrodes. Alternate channel combinations were not explored due to time constraints.

Both participants donned myoelectric prostheses to perform the SHAP-inspired tasks. Participants relied solely on their robotic prostheses for visual feedback of the pattern recognition system. Durplex sockets were fabricated for each participant’s residual limb at the University of Michigan Orthotics and Prosthetics center by a certified prosthetist. To control physical prostheses, the xPC sent commands via serial connection to a custom circuit board which issued the appropriate CAN messages for each hand. The software interface controlled robotic prostheses by directly translating predicted postures to closed grasps while rest commands opened the hand. For P1, the internal controller on the LUKE arm operated in position control mode. Finger positions corresponding to the endpoints of the current predicted posture were sent to the LUKE arm, which processed updates every 10ms. For P2, a state machine was coded to interface between the xPC and i-Limb which closed and opened selected grips. The i-Limb interface introduced a hardware delay due to the time to process a grip change. However, the compact form factor of the extra small i-Limb Quantum was greatly preferred for P2, who could not lift the LUKE arm. The SHAP-inspired task was evaluated by trial time and two qualitative metrics. A sub-task was counted as complete if it was performed in a single attempt. It was possible to complete tasks without using the instructed grasp. The number of sub-tasks where the correct grasp was achieved without multiple activation attempts was also counted.

## Data Availability

Data used in this study may be made available upon reasonable request.

## Acknowledgments

The authors would like to acknowledge Christina Lee for collecting range of motion measurements and assisting during experiments, as well as our clinical trials coordinator Kelsey Ebbs for managing regulatory compliance. We would also like to thank our colleagues Michael Gonzalez for building a socket adapter for the LUKE arm, and finally John Busch for assistance developing the RS232-CAN interface for the LUKE arm.

## Funding

This work was supported by the Defense Advanced Research Projects Agency (DARPA) Biological Technologies Office (BTO) Hand Proprioception and Touch Interfaces (HAPTIX) program through the DARPA Contracts Management Office grant/contract no. N66001-16-1-4006 and by the National Institute Of Neurological Disorders And Stroke of the National Institutes of Health under Award Number R01NS105132 to C.A.C. P.P.V. was supported by the National Science Foundation Graduate Research Fellowship Program under Award Number DGE 1256260. The opinions expressed in this article are the authors own and do not reflect the view of the Department of Defense, National Institutes of Health, or the National Science Foundation.

## Author contributions

A.K.V., P.P.V., D.H.G., and C.A.C. designed the study. A.K.V., P.P.V., N.N., and A.J.D. completed the data gathering and development of analysis techniques with all authors contributing to the interpretation of results. P.S.C. and T.A.K. developed and performed the surgical implantation procedures. A.K.V. and P.P.V. developed the software for the real-time experiments. A.J.D. fabricated and fit the prototype sockets. All authors contributed to writing, critical review, and approval of the report.

## Competing interests

The University of Michigan holds a patent related to this work, publication number US20190262145A1: System for amplifying signals from individual nerve fascicles.

## Data and materials availability

All processed data associated with this study are present in the paper or the Supplementary Materials. Raw data may be made available upon reasonable request.

## Notes

### Competing Interest Statement

The authors have declared no competing interest.

### Clinical Trial

NCT03260400

### Author Declarations

All experiments were conducted in accordance with the University of Michigan Institutional Review Board, study ID HUM00124839. For both participants, informed consent was obtained after the nature and possible consequences of the study was explained. Both participants also signed consent forms for media deidentification.

