## Supplementary Materials for "Surgically Implanted Electrodes Enable Real-Time Finger and Grasp Pattern Recognition for Prosthetic Hands"

### Materials and Methods

#### *Hidden Markov Model Performance Analysis*

The HMM-NB was compared to a standard Naïve Bayes (NB) classifier using the virtual task to switch between postures for the 1 of 10 and grasps sets (Table S1). Parameters for each decoder are shown in Table S3. For both participants, classifier parameters were selected by completing preliminary real-time control tests with larger window sizes and more conservative filtering and decreasing both parameters until instabilities prevented task completion. The output for the HMM-NB at each time step was updated if the sum of probabilities of hold or rest states exceeded 0.8. No additional output filters were applied to the HMM-NB, with the exception of P1's implementations with 10 ms updates. Both the NB and HMM used the same calibration data in each session. P1 completed one A-B session with the NB (A) and HMM-NB (B) classifiers for the 1 of 10 and Grasps posture sets. P2 completed three sessions for posture set: one A-B, one A-B-A, and one B-A-B session for 1 of 10 and three A-B-A sessions for Grasps. Fig. S2 and Fig. S3 show the results of the NB classifier which can be compared to the HMM-NB from Fig. 1 and Fig. 2. P1 completed the 1 of 10 posture set faster with the HMM-NB, achieving a latency of 159±237 ms compared to 258±313 ms (median±i.q.r.). However, the NB was more stable evidenced by a higher online accuracy, 95.9% compared to 93.0%, and fewer trials with a latency greater than one second. The HMM-NB outperformed NB in both online metrics for Grasps. NB achieved a high offline Grasp accuracy for P1, but had a lower online accuracy of 83.7% compared to 99.5%. The increase in transition errors contributed to a higher median latency of 280±251 ms compared to 96±30 ms for the HMM-NB. For P2, the HMM improved performance of both posture sets. For 1 of 10, P2's NB classifier had a median latency of 398±1246 ms compared to 344±924 ms with the HMM-NB. Transition errors were worse overall as she achieved an online accuracy of 75.7% with NB compared to 80.0% with the HMM-NB. However, the NB classifier was able to return to rest more efficiently than the HMM-NB. For the Grasps posture set, the NB classifier was far less responsive with a latency of 355±1184 ms compared to 173±151 ms. Transition errors were also much worse with an online accuracy of 81.5% compared to 96.3% with the HMM-NB.

The HMM-NB particularly excelled in distinguishing the Grasp posture set. The ability to represent a posture as multiple states could be a greater advantage for predicting compound finger movements. The simplicity of the Naive Bayes assumption within the HMM-NB meant that latent states could not represent complex phenomena, but also meant the model was not as prone to over-fitting as more powerful techniques (*63*). The incomplete selection of latent states for output was problematic for P2's 1 of 10 decoders which sometimes got stuck in transition states while returning to rest. Supervised learning with alternate output mappings could be explored to mitigate this issue. Expectation-Maximization algorithms can settle into local minima. Here, the low amount of training data, 5-7 movement repetitions, meant the HMM-NB was especially sensitive to state structure and initialization. Therefore, we cannot say the specific implementation chosen here was the best model, rather one that worked well. The number and structure of latent states per posture was driven by the computational requirements of the real-time system, ease of initialization, and success in prior applications (*40, 64*). Relaxing the state structure and initialization routine could allow the HMM-NB to converge to more optimal solutions, although this would likely require more training data. As noted elsewhere, assembling large training data-sets is feasible due to the signal stability of implanted electrodes over time, which has been documented in other work (*31*, *38)*.

*Offline Analysis and Signal Comparisons*

The 1 of 10 posture set was chosen for alternate classifier simulations because it required classifiers to distinguish between the most movements. It also requires distinction between individual fingers, some of which were well represented by electrode placement and some of which were not. The algorithms used for real-time control were compared offline to a Naive Bayes (NB), linear discriminant analysis (LDA), and a multi-class support vector machine (SVM) using five time domain features (mean absolute value, waveform length, variance, slope sign changes, and zero crossings) along with coefficients from a $6^{th}$ order auto-regressive model. Alternate classifiers assumed equal prior probabilities for each posture and used default Matlab 2018 built-ins for training and evaluation. LDA used a diagonal-regularized pooled covariance matrix. The multi-class SVM used a one-vs-one architecture with a linear kernel. Performance was evaluated on calibration data with different sized processing windows containing 10, 25, 50, 100, 150, 200, and 250ms of EMG history. P1's HMM-NB for comparison was updated in 10ms timesteps matching his online implementation (Table S3). All other classifiers were updated every 50ms for lengthier processing windows.

P2 completed additional sessions to compare the performance between implanted and surface EMG. For the classifier simulation, eight pairs of adhesive electrodes were placed on P2's residual limb. Surface muscles corresponding to implanted electrode functions were targeted by feeling P2's forearm while asking her to perform movements with her phantom limb. The size of adhesive electrodes resulted in the majority of her medial forearm being covered. EMG from both the surface and implanted electrodes was simultaneously recorded while P2 performed a calibration run for the 1 of 10 posture set. In a separate session we also precisely targeted FCR, FDPS, FPL and EPL using established techniques (*65*). For targeted sessions, recordings were done individually to avoid space constraints. Before each recording, P2's forearm was cleaned with alcohol wipes and allowed to dry before applying the gelled electrodes. Signals from the corresponding implanted electrode pair were also recorded for the simultaneous comparison. The same calibration routine was used to instruct movement cues that corresponded to the muscles' motor functions. SNR's were calculated by averaging the RMS voltage of active periods and dividing by the averaged RMS of rest periods. Rest periods sometimes began with EMG settling activity from the previous trial. This was particularly noticeable for some of P2's surface channels as well as her FDPI and EPL implanted electrodes. SNR comparisons were conducted for surface and implanted channels targeting FDPI, EPL, and FCR. The SNR of RPNI electrodes was roughly compared to surface by targeting FPL and FDPS, residual muscles with similar motor functions. SNR analysis was performed on both the targeted and classifier calibration datasets. Settling activity was manually removed for all SNR analysis, but not for classifier training because it is important to the characterization of a rest intention. For each movement and muscle pair, the session with the better surface SNR was chosen for presentation in Fig. 6. For FCR and FPDS, the targeted sessions yielded better results, while EPL, FDPI, and FPL comparisons were taken from the classifier calibration dataset.

*Participant Anatomy and Experiment Set-Up*

RPNIs are created by suturing a small muscle graft to the end of severed nerve. In addition to preventing neuroma growth, RPNIs serve as bioamplifier for afferent motor signals and have been demonstrated to produce stable functionally selective EMG in animal and human studies (*35,36,38*). P1 is male in his 30's who sustained a right wrist disarticulation. In 2015 he underwent surgery to resect neuromas on the median, ulnar, and radial nerves. A single RPNI was created on each nerve using free skeletal muscle grafts from his ipsilateral vastus lateralis. P1 is not a body powered user. In 2018, P1 underwent an additional surgery to have eight pairs of bipolar intramuscular electrodes (Synapse Biomedical, Oberlin, OH) chronically implanted into the median and ulnar RPNIs and residual muscle corresponding to hand and wrist functions. P1 had the following residual muscles targeted for implantation: Flexor Pollicis Longus (FPL), Flexor Digitorum Profundus - Index Section (FDPI), Flexor Digitorum Profundus - Small Section (FDPS), Extensor Pollicis Longus (EPL), Extensor Digitorum Communis (EDC), and Flexor Carpi Radialis (FCR). P2 completed experiment sessions for this study from 2018 to 2019. The electrodes remained implanted for approximately one year and were partially explanted in 2019 by removing any exposed and subcutaneous wire lengths not embedded in muscle. In 2020, P1 had his electrodes fully explanted from RPNIs and residual muscles.

P2 is a female in her 50's who underwent a voluntary transradial amputation in 2017. Single RPNIs were created on each of the median and radial nerves while an intraneural dissection was performed on the ulnar nerve to create two RPNIs. All RPNIs were creating using free muscle grafts from the ipsilateral vastus lateralis. P2 currently uses a body powered prostheses outside of the study, although she reports to seldom use the open-close functionality. In 2018, P2 underwent chronic implantation of eight pairs of bipolar electrodes into the median and both ulnar RPNIs and five residual muscles. The same residual muscles were targeted for both patients with the exception of FDPS, which was only targeted for P1. P2 completed experiments for this study from 2019 to 2020. At the time of writing, P2 remains implanted. In 2017 P2's right passive elbow range of motion was recorded in clinic to be 20 – 120° of flexion. Clinicians also noted limited ability to supinate her forearm with maximal supination in neutral at 0°. In 2020, experimenters measured her passive right shoulder range of motion to be 160° shoulder flexion and 90° external rotation. We measured shoulder external rotation with the participant lying on her back, arm abducted to 90°, shoulder flexed to 90°, and forearm prone. In 2020, her elbow range of motion was again measured by experimenters to be 20 – 125° of flexion.

The experiment set-up also differed slightly between subjects. P1 completed calibration runs with pseudo-randomly ordered cues and rest and hold periods of 2.5 seconds each. P2 preferred a slower pace and performed the calibration run with rest and hold periods of 3 seconds each and consecutively ordered cues. For the grasps posture set, P1's visual prompt showed finger extension instead of finger abduction. In preliminary calibration sessions, we found these two cues produced similar EMG responses. P1's prompt for point also resembled small finger flexion. This cue was consistent with other physical prostheses experiments P1 performed with the LUKE arm, which can only move its small finger in combination with the middle and ring fingers. A custom adapter was made to connect the LUKE arm to P1's socket, which was secured to his forearm with an Otto Bock silicone liner (Ottobock, Duderstadt, Germany). His socket also featured a window to allow access to the percutaneous electrode connectors on his medial forearm. For P2, the i-Limb was connected to her socket with a quick wrist disconnect (QWD) that allowed manual wrist rotation. The QWD was embedded in a PVC adapter that connected to her socket which was secured to her forearm with an Iceross Upper-X liner (Ossur, Reykjavik, Iceland). Her percutaneous leads exited on her lateral bicep and did not interfere with her socket.

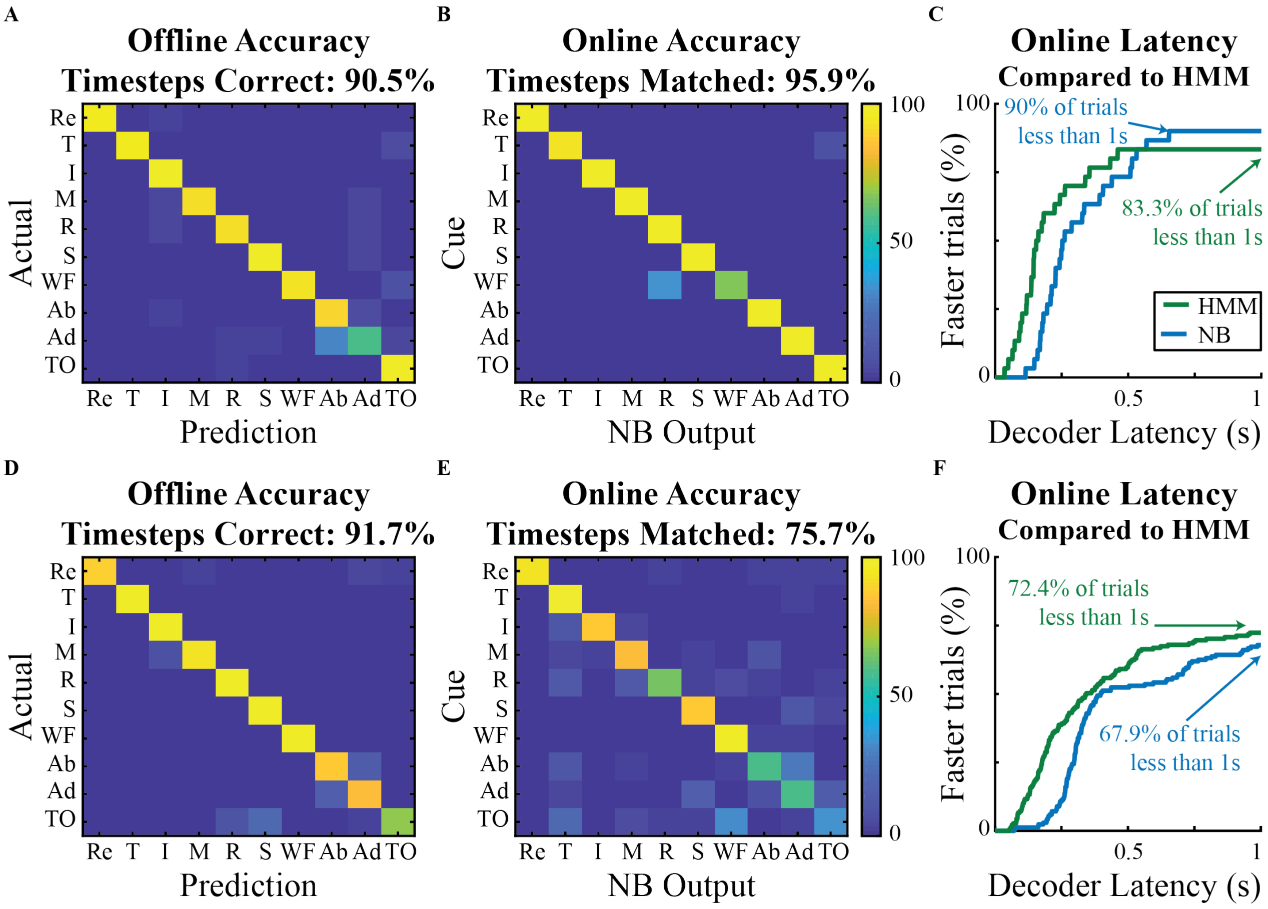


**Fig. S1. Performance of the Naive Bayes classifier on the 1 of 10 posture set. (A)** Offline classifier accuracy simulated on individual time bins during hold periods using 5-fold cross validation on P1's training data. **(B)** An output filter of three consecutive decodes was used for real-time control and P1 was able to achieve an online accuracy of 95.9%. **(C)** Cumulative latency lines are drawn so the y-axis indicates the percentage of trials with latency less than values on the x-axis (n = 30 trials, 27 shown). Results compared to the HMM results from Fig. 1 **(D,E)** P2's decoder used larger processing windows and a filter length of four consecutive decodes for real-time control. **(F)** Cumulative latency comparison for P2 (n = 168 trials, 114 shown).

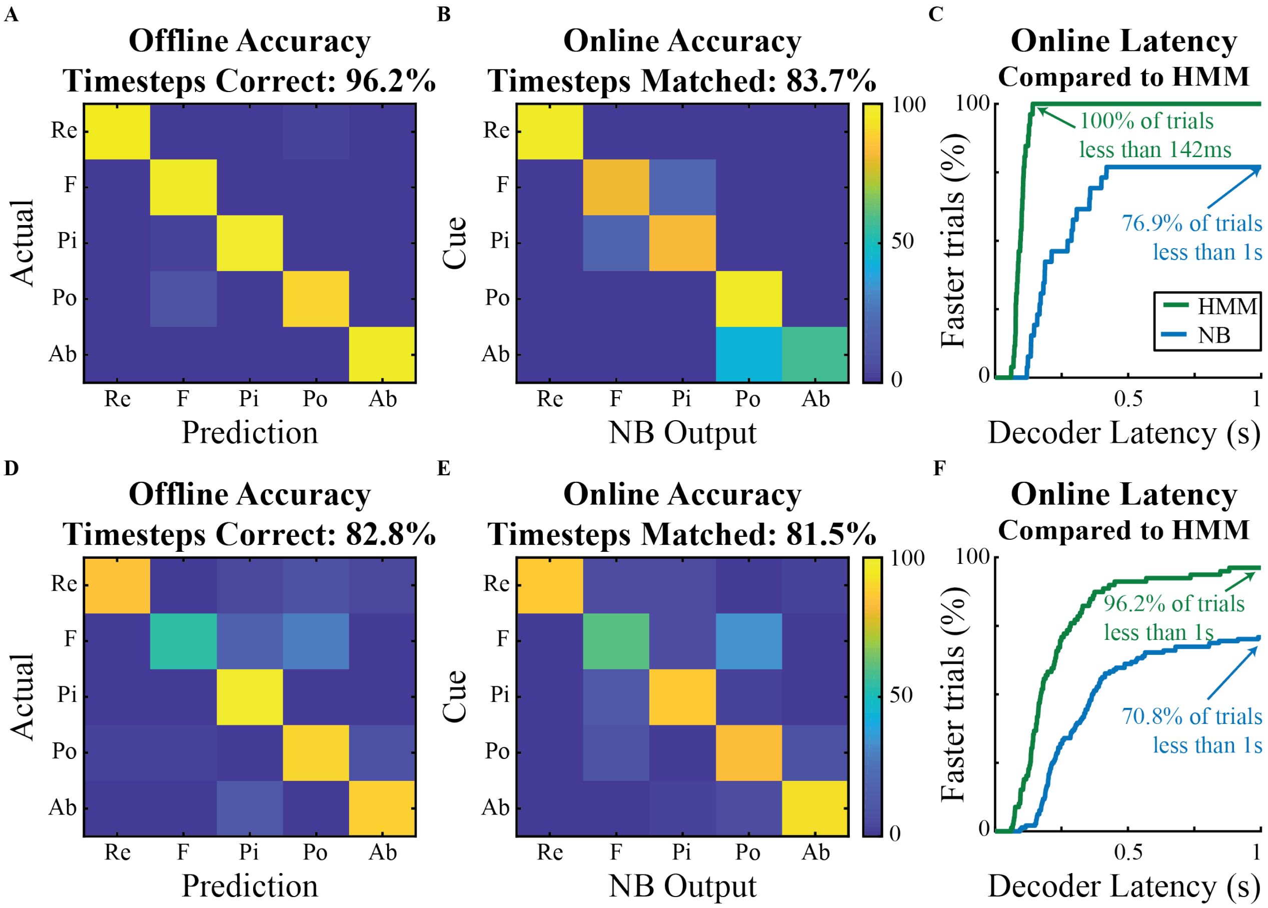


**Fig. S2. Performance of the Naive Bayes classifier on the Grasps posture set. (A)** Offline classifier accuracy simulated on P1's training data. **(B)** An output filter of three consecutive decodes was again used for real-time control. **(C)** Cumulative latency comparison for P1 (n = 26 trials, 20 shown). Results compared to the HMM from Fig. 2. **(D,E)** P2's decoder again used larger processing windows and a filter length of four consecutive decodes for real-time control. **(F)** Cumulative latency comparison for P2 (n = 144 trials, 102 shown).

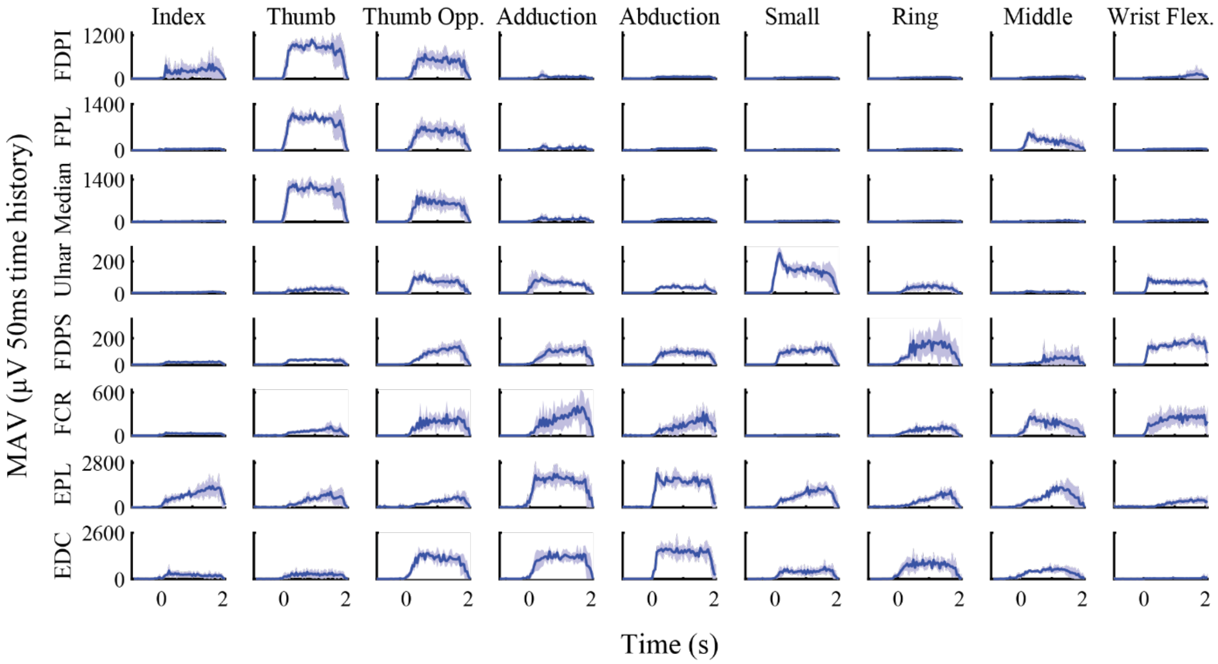


**Fig. S3. Channel activity from P1's calibration session.** Mean absolute value (MAV) traces of P1's EMG during the 1 of 10 posture set used for classifier training and clustering analysis. EMG was rectified and averaged in non-overlapping 50ms time bins from electrodes targeting flexor digitorum profundus, index section (FDPI), flexor pollicis longus (FPL), Median RPNI, Ulnar RPNI, flexor digitorum profundus, small section (FDPS), flexor carpi radialis (FCR), extensor pollicis longus (EPL), and extensor digitorum communis (EDC). Trials were time aligned to the start of the hold period and then averaged (mean±s.t.d. n = 5-6 trials).

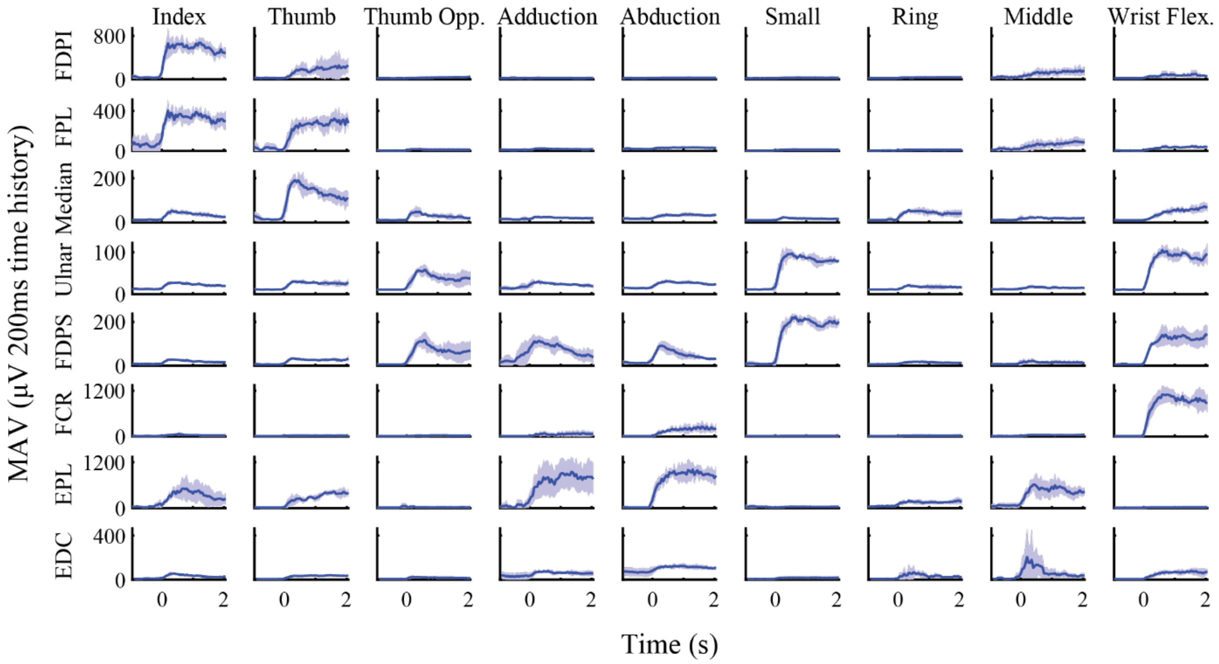


**Fig. S4. Channel activity from one of P2's calibration sessions.** Mean absolute value (MAV) traces of P2's EMG during the 1 of 10 posture set from the experiment session used for offline and clustering analysis. EMG was rectified and averaged in 200ms time bins with a 50ms update rate. Electrodes targeted flexor digitorum profundus, index section (FDPI), flexor pollicis longus (FPL), Median RPNI, Ulnar RPNI 1, Ulnar RPNI 2, flexor carpi radialis (FCR), extensor pollicis longus (EPL), and extensor digitorum communis (EDC). Trials were time aligned to the start of the hold period and then averaged (mean±s.t.d, n = five trials).

| Study | Participants | EMG Channels | Hand Classes | Completion Delay (s) | Completion Rate (%) |
|---|---|---|---|---|---|
| Kuiken 2009 (*27*) | 3 SD, 2 TR | 12 surface gelled | 4 | 0.54±0.27 | 86.9±13.9 |
| Li 2010 (*14*) | 5 TR | 12 surface gelled | 6 | 0.45±0.35 | 53.9±14.2 |
| Cipriani 2011 (*18*) | 5 TR | 8 surface gelled | 7 | 0.86* | 77* |
| Vaskov et al. 2020 | 2 TR | 8 intra-muscular | 8 | 0.26±0.09 | 96.3±5.3 |
| | | | 4 | 0.14±0.06 | 100±0.0 |

**Table S1. HMM comparison to previous work.** Three earlier studies quantified real-time classification including multiple hand (finger or grasp) movements for persons with shoulder disarticulations (SD), transhumeral (TH), or transradial (TR) amputations using similar randomized control tasks. Movement completion metrics for hand postures were calculated consistent with previous work. The task in those studies differed from the posture switching task in two ways: a rest period was presented in between cues and the requirement for completion was a cumulative selection rather than a continuous hold. Completion rate and time were defined as the percentage of trials and median time in which one second of the correct posture was cumulatively matched. Completion delay is presented as the difference between the reported completion time and the cumulative selection length, which differed amongst earlier studies. By nature, completion rate is greater than or equal to success rate and completion delay is less than or equal to latency. Metrics were averaged across subjects (mean±s.t.d.) *variance across subjects not reported.

| Posture Set | No. of Classes | Classes |
|---|---|---|
| 1 of 10 | 10 | Thumb, Index, Middle, Ring, and Small Finger Flexion (T,I,M,R,S)<br>Wrist Flexion (WF), Finger Abduction (Ab), Finger Adduction (Ad), Thumb Opposition (TO), Rest (Re) |
| Grasps | 5 | Fist (F), Pinch (Pi), Point (Po),<br>Finger Abduction (Ab), Rest (Re) |
| Functional | 4 | Fist (F), Pinch (Pi), Point (Po), Rest (Re) |

**Table S2. Posture sets uses in the study.** P2 performed three experiment sessions of the virtual task with the 1 of 10 and grasps posture sets, while P1 performed one session with each set. The functional posture set was used by both participants to control robotic prostheses and for the static arm position test performed by P2.

| Participant | Task | Decoder | Time History (ms) | Update Rate (ms) | Filter Length |
|---|---|---|---|---|---|
| P1 | 1 of 10 | HMM | 50 | 10 | 5 |
| | | NB | 50 | 50 | 3 |
| | Grasps | HMM | 50 | 10 | 5 |
| | | NB | 50 | 50 | 3 |
| P2 | 1 of 10 | HMM | 50 | 50 | 1 |
| | | NB | 200 | 50 | 4 |
| | Grasps | HMM | 50 | 50 | 1 |
| | | NB | 200 | 50 | 4 |

**Table S3. Decoder parameters for real-time control comparison**. Time history refers to the length of the processing window to extract MAV from all eight bipolar electrode pairs, while update rate refers to the timestep features and decoders were updated. The filter length is the number of consecutive decodes required for a change to be sent to the virtual prostheses.
